# Understanding Vaccination Uptake amongst Gay, Bisexual and other Men who have Sex with Men in UK Sexual Health Services: A Qualitative Interview Study

**DOI:** 10.64898/2025.12.22.25342671

**Authors:** Tom May, Joanna M. Kesten, Hannah E. Family, Harriet Fisher, Adele Wolujewicz, Marta Checchi, Hamish Mohammed, David Leeman, Sema Mandal, Lucy Yardley, Jeremy Horwood, Clare Thomas

**Affiliations:** National Institute of Health and Care Research Health Protection Research Unit in Evaluation and Behavioural Science at the University of Bristol, Bristol, UK; National Institute for Health and Care Research Applied Research Collaboration West at University Hospitals Bristol and Weston NHS Foundation Trust, Bristol, UK; Yuno Sexual Health, University Hospitals Bristol and Weston NHS Foundation Trust, Bristol, UK; Blood Safety, Hepatitis, STI and HIV Division, UK Health Security Agency, London, UK; School of Psychological Science, University of Bristol and School of Psychology, University of Southampton; Centre for Academic Primary Care, Bristol Medical School, University of Bristol, Bristol, UK

**Keywords:** Vaccination, Human papillomavirus, hepatitis A virus, hepatitis B virus, Sexual Health, GBMSM

## Abstract

**Background/Objectives:** In England, gay, bisexual and other men-who-have-sex-with-men (GBMSM) are eligible for vaccinations at NHS sexual health services, including human papillomavirus (HPV), hepatitis A virus (HAV), and hepatitis B virus (HBV). However, current research shows limited understanding of the factors influencing vaccination uptake among GBMSM. This study aimed to examine the barriers and facilitators affecting the offer and uptake of these vaccination programmes.

**Methods:** A qualitative interview study following the Person-Based Approach (a systematic method for developing and optimising health interventions) involving GBMSM and sexual health service staff from two regions of England. Purposive sampling aimed to include GBMSM with diverse backgrounds and engagement with sexual health services. Patient and public involvement shaped the study design and interview topic guides. The interviews were recorded, transcribed, and thematically analysed to identify barriers and facilitators which were interpreted using the COM-B model of behaviour change.

**Results:** Twenty GBMSM and eleven staff took part. The findings showed that opportunistic delivery of HPV, HAV and HBV vaccination within sexual health services is mostly acceptable and feasible for GBMSM and staff while also highlighting areas for optimization. Despite low knowledge of these viruses and their associated risks, willingness to be vaccinated was high, with healthcare provider recommendations and the convenience of vaccine delivery during routine clinic visits acting as important facilitators. However, the reach of opportunistic models was limited, particularly for individuals underserved by sexual health services or disengaged from GBMSM social networks. System-level barriers such as complex vaccine schedules (particularly when multiple schedules are combined), inconsistent access to vaccination histories and limited system-level follow-up processes (e.g. automated invites and reminders) were also found to act as obstacles to vaccination uptake and delivery.

**Conclusions:** To improve equitable uptake, sexual health services should explore the feasibility of addressing both individual and structural barriers through additional strategies, including targeted and persuasive communication to increase knowledge, leveraging regular contact with GBMSM to promote uptake and implementing enhanced approaches to support vaccination completion (e.g. prompts or reminders).

## 1. Introduction

Gay, bisexual, and other men who have sex with men (GBMSM) are at greater risk of human papillomavirus (HPV), hepatitis A virus (HAV) and hepatitis B virus (HBV) infections which can be transmitted through sexual contact (1). Persistent infection with HPV can lead to the development of anogenital warts and some cancers. HBV infection can become chronic and lead to liver damage, cancer or failure, whilst HAV infection is acute and does not become chronic, but comorbid conditions (including HBV, hepatitis C and HIV) may increase the risk of liver failure following infection (5). Effective vaccines for the prevention of these infections are available (5–8). In England, vaccines are available free of charge through national health service (NHS) specialist sexual health services for those who meet eligibility criteria, including GBMSM (5–8)^1^. The commissioning and procurement pathways, however, vary for the different vaccines and changes to vaccine schedules, including the addition of new vaccination programmes (such as those against Mpox and gonorrhoea), add complexity to programme delivery.

Research suggests GBMSM generally hold positive views about vaccination (9). However, estimates of uptake rates vary considerably. For example, a UK community survey found self-reported uptake rates of 68-73% (10), but levels of HPV vaccine uptake reported using clinical electronic patient record data were much lower at 45- 49% (11, 12). More recent data for England indicates that cumulative HPV vaccine uptake to the end of 2024 was 32–49% (13). Known issues with under-reporting in clinical records, under-representation in younger age groups and non-white ethnicities in the survey sample and difficulties in reliably confirming immunity from verbal vaccine history, particularly when vaccinations received in different settings are not recorded within sexual health service records to protect confidentiality, also means there is still uncertainty about the true levels of vaccine coverage. Similar issues persist for HBV vaccination coverage among GBMSM, where incomplete coding of immunity or vaccination in sexual health services may contribute to inaccurately low estimates of vaccination coverage (14). In addition, while the Joint Committee on Vaccination and Immunisation (JCVI) recommends targeting vaccination to GBMSM attending sexual health services due to their higher risk, reliance on opportunistic delivery within these settings may limit broader coverage, given that only 33% of GBMSM reported attending a sexual health clinic in the last year (15).

Previous research has found that multiple interacting factors influence GBMSM vaccination uptake in sexual health services, relating to the individual, healthcare provider, organisational setting, and community and policy environments (9). Further, HPV vaccine uptake amongst GBMSM was found to be influenced by the limited perceived relevancy of HPV, often due to lack of information and the feminisation of HPV (e.g. promotion of the link between HPV and cervical cancer), as well as sociocultural contexts and healthcare experiences. The latter encompassed the influence of the healthcare provider recommendations of vaccination and the role of disclosure of GBMSM identity to indicate eligibility for vaccination (16). For healthcare professionals, time pressure in consultations, difficulty identifying eligible GBMSM and not feeling sufficiently informed were identified as barriers to vaccine delivery (17–19).

Interventions found to be successful in increasing uptake amongst GBMSM include the provision of information combined with motivational messaging, and interaction with or recommendation by healthcare professionals (16). In addition, the use of telephone contact, text messaging and online communication were all found to be acceptable ways to receive information (20–25). Outreach health promotion activities such as offering vaccinations at annual LGBTQ+ Pride events were found to be effective at targeting populations who may not access sexual health services, as well as GBMSM who are engaged in high-risk sexual behaviours (26, 27).

However, the existing literature has limitations as most studies focus on uptake of single vaccines rather than considering them in combination, and the majority were conducted prior to the COVID-19 pandemic. As a result, it is important to explore whether COVID-19–related vaccine fatigue (28) may exist among GBMSM, and how this might influence acceptance of current vaccination programmes. This is particularly important in the context of recent shifts towards the online delivery of some sexual health services in England - such as self-sampling testing for sexually transmitted infections (STIs) - which may reduce the need for in-person clinic attendance. Although previous research has indicated both GBMSM and healthcare professionals would welcome more information or training on HPV, HAV and HBV infections and vaccines (19, 29), framing findings with reference to behaviour change theories would also help to better understand how information provision can be used most effectively to increase vaccination uptake and identify any additional interventions required.

### Study Aims and Objectives

The overall aim of this study was to explore ways to increase the uptake of HPV, HAV and HBV vaccination programmes amongst GBMSM, informed by behaviour change theory. The specific objectives were to:

1. Explore the attitudes amongst GBMSM from diverse backgrounds towards HPV, HAV and HBV infections and vaccines, and identify barriers and facilitators to vaccine uptake.
2. Explore the views of sexual health service staff and commissioners on the delivery of vaccines to GBMSM and identify barriers and facilitators to vaccine programme delivery post COVID-19 pandemic.
3. Develop recommendations based on the study’s findings to optimise service delivery and improve information provision to staff and GBMSM with the aim of increasing the offer and uptake of HPV/HAV/HBV vaccination programmes.

## 2. Materials and Methods

### 2.1. Design

A qualitative study, employing semi-structured interviews by telephone or online. The study methodology was informed by the Person-Based Approach (PBA) (30), which focuses on developing and optimizing interventions to ensure they are tailored to the views, needs and experiences of individuals who use them. PBA uses an iterative approach to intervention planning and optimisation, incorporating input from patient and public involvement and qualitative research.

PBA has two broad areas of activity: 1) Gathering evidence about user context and experience, and 2) Programme theory and intervention development (31). This manuscript reports a qualitative study conducted within the first of these two areas of activity, which supports the second by identifying likely intervention mechanisms to inform programme theory. This process also included conducting a review of relevant literature, scoping conversations with GBMSM public contributors and sexual health clinicians, and systematically recording qualitative findings, including barriers and facilitators to target behaviours (intervention uptake and delivery) in an Intervention Planning Table^2^. Findings were also discussed iteratively throughout the data collection and analysis period with the study team, consisting of Blood Safety, Hepatitis, STI and HIV Division colleagues from the UK Health Security Agency (UKHSA), a sexual health consultant and behavioural science experts.

### 2.2. Setting

The study took place in South-West and North-West England between August 2024 and February 2025 and was supported by two sexual health services operating in these regions. At the time of the study, the new national vaccination programmes for Mpox and 4CMenB for gonorrhoea were not yet available in these services.

### 2.3. Participants, Recruitment and data collection procedures

GBMSM participants were recruited either through the two participating sexual health services or by using wider community recruitment strategies. In the sexual health services, eligible GBMSM were given verbal information and printed participant information sheets by a member of the healthcare team during a clinical consultation. Recruitment posters were also displayed in the waiting rooms. To ensure the study included the perspectives of GBMSM who were not currently engaging with sexual health services, the study information and a recruitment poster were promoted by local community organisations and representatives who work with GBMSM. This included distribution through local community WhatsApp groups and social media. Inclusion criteria were: GBMSM, based in either of the two study locations, 18 years old or above, and able to provide informed consent.

Individuals expressed their interest in taking part on an online sign-up page which recorded contact details and basic demographic information. Recruitment was purposive, based on age, vaccination status, ethnicity, socioeconomic background, and engagement (or not) with sexual health services. A sample size of approximately 25 GBMSM participants was planned, based on our previous experience of undertaking qualitative studies (32) and a pragmatic assessment of the numbers required to obtain sufficient data according to the concept of information power (33).

Sexual health service commissioners and staff with different job roles (e.g. doctors, nurses, public health practitioners and data managers) and levels of experience involved in the delivery of the HPV, HAV and HBV vaccination programmes were recruited from the two participating sexual health services and through existing connections with commissioning organisations. The lead clinician in each service facilitated invitation distribution by sharing information sheets and consent forms with their colleagues, who then contacted the research team if they wished to participate. All participants were made aware of the incentive for participating (£30 shopping e-voucher).

GBMSM and staff who expressed interest in taking part were contacted by the research team to discuss what participation would involve, re-provide the participant information sheet, and agree a convenient time for the interview. The interviews were semi-structured, guided by topic guides which covered knowledge of the vaccination programme and the diseases protected against, barriers and facilitators to uptake and delivery and suggestions for ways to improve provision (see supplementary information). The topic guides were developed with reference to scoping conversations between the research team and GBMSM public contributors and sexual health clinicians. The interviews took place either on Microsoft Teams or by telephone. Follow-up telephone calls, texts or email to non-responders were used up to three times as required. Written consent was provided by participants ahead of the interview using an online form via a link which was emailed or texted to them, along with an invitation to email with questions or talk through the form on a call. Before the interview started, participants were briefed again on the purpose for the interview and key information about the study and the right to withdraw, before giving verbal consent for the interview recording to begin. Interviews were audio recorded using a digital encrypted Dictaphone, or the record function in Microsoft Teams. At the end of the interview, participants were thanked and provided with a £30 shopping e-voucher.

### 2.4. Data Analysis

The interviews were transcribed verbatim and then analysed inductively and deductively to identify barriers and facilitators to the offer and uptake of vaccination. TM and HEF analysed the GBMSM data and JK analysed the staff data. We used the Intervention Planning Table to detail the barriers and facilitators to the target behaviours (GBMSM taking up vaccination offer & sexual health services offering vaccinations) identified in the interviews. The interview analyses were reviewed across staff and GBMSM transcripts and interpreted using the Capability, Opportunity, Motivation - Behaviour (COM-B) (34) model of behaviour change to create a “behavioural diagnosis” of the barriers and facilitators to the target behaviours. The model proposes that in order for a behaviour to occur, an individual must have three things: (1) capability to carry out the behaviour, including physical capability (e.g. skills) and psychological capability (e.g. knowledge, memory); (2) opportunity to carry out the behaviour including social opportunity (e.g. the behaviour fits with social norms) and physical opportunity (e.g. the person has the time, resources and can access what is needed to carry out the behaviour); and (3) motivation to undertake the behaviour including automatic motivation (which is influenced by emotional responses or habit) and reflective motivation (confidence they can carry out the behaviour, belief that the behaviour will be beneficial for them). The COM-B behavioural diagnosis can then be used to facilitate selection of evidence-based behaviour change techniques to enhance facilitators and address barriers. Although factors could fit within multiple COM-B components, we have primarily categorised them based on how they functioned to drive behaviour, for example, by facilitating opportunities for action, increasing motivation or supporting capabilities to perform a behaviour (35).

## 3. Results

We interviewed 20 GBMSM, with equal numbers across our study sites in the South-West and North-West of England. Participants ranged in age from 20 to 64 years. Eight participants identified as non-white. Four participants self-reported that they had not received any of the three vaccines. Sixteen participants reported that they had received at least one of the three vaccines, whilst eight participants reported that they had all three vaccines. There was also variation in the setting these vaccines had been received, including as part of the universal schools-based HPV vaccination programme and vaccination for HAV from travel clinics.

Some participants expressed uncertainty about which vaccines they had previously received. This uncertainty partly reflected the absence of a consolidated vaccination record and the inability to view sexual health vaccination history via the NHS App, as sexual health services maintain separate records to protect patient confidentiality across services (36).

11 staff completed interviews from the participating sexual health services, five from the service in South-West England and six from the service in North-West England. Staff had diverse roles including clinical, data management and public health commissioning.

Further information about participant characteristics is provided in Table 1.

**Table 1.**
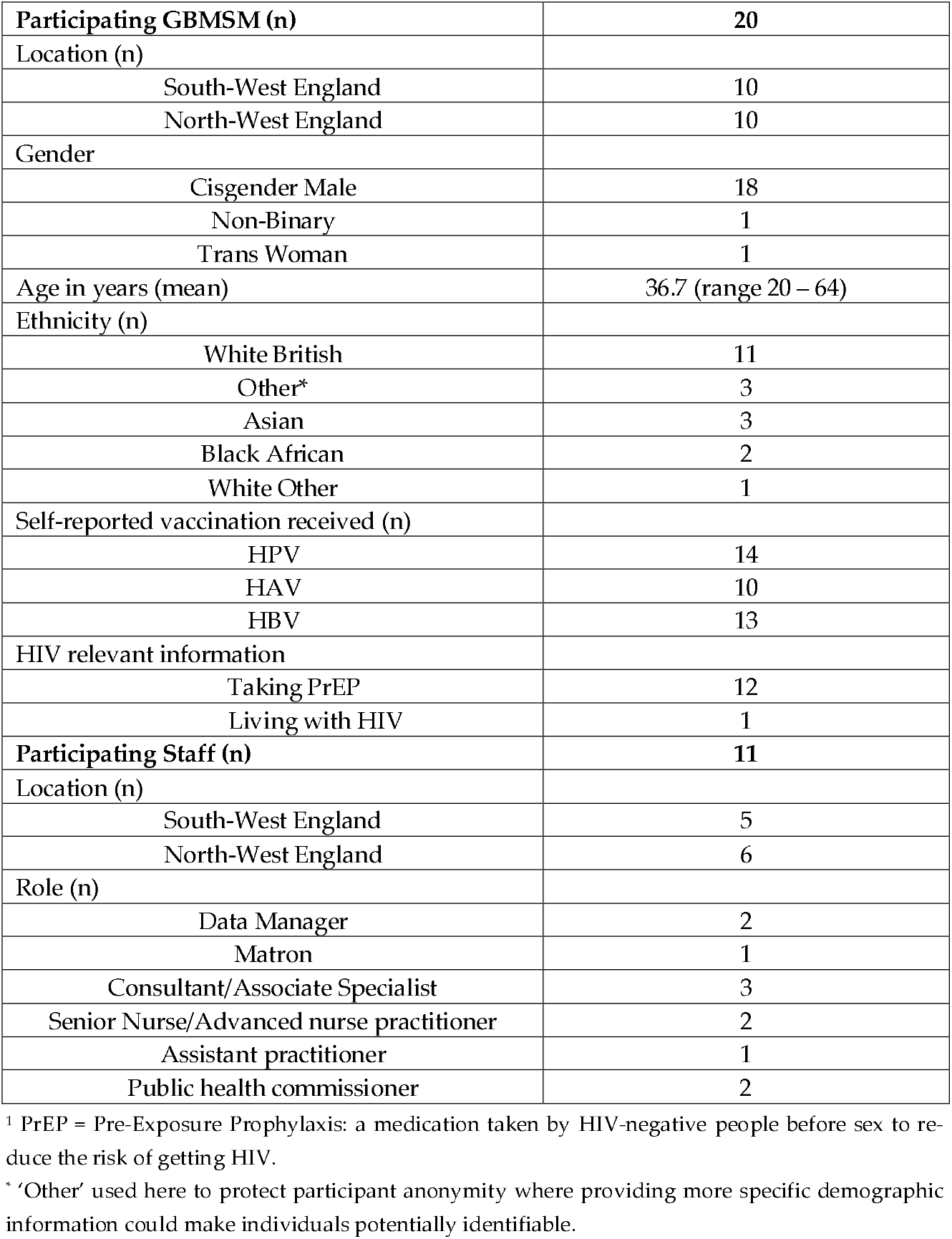
Participant Characteristics.

|  |  |
| --- | --- |
| **Participating GBMSM (n)** | **20** |
| Location (n) |  |
| South-West England | 10 |
| North-West England | 10 |
| Gender |  |
| Cisgender Male | 18 |
| Non-Binary | 1 |
| Trans Woman | 1 |
| Age in years (mean) | 36.7 (range 20 – 64) |
| Ethnicity (n) |  |
| White British | 11 |
| Other* | 3 |
| Asian | 3 |
| Black African | 2 |
| White Other | 1 |
| Self-reported vaccination received (n) |  |
| HPV | 14 |
| HAV | 10 |
| HBV | 13 |
| HIV relevant information |  |
| Taking PrEP | 12 |
| Living with HIV | 1 |
| **Participating Staff (n)** | **11** |
| Location (n) |  |
| South-West England | 5 |
| North-West England | 6 |
| Role (n) |  |
| Data Manager | 2 |
| Matron | 1 |
| Consultant/Associate Specialist | 3 |
| Senior Nurse/Advanced nurse practitioner | 2 |
| Assistant practitioner | 1 |
| Public health commissioner | 2 |

<sup>1</sup> PrEP = Pre-Exposure Prophylaxis: a medication taken by HIV-negative people before sex to reduce the risk of getting HIV.
\* 'Other' used here to protect participant anonymity where providing more specific demographic information could make individuals potentially identifiable.

Analysis of the interview data identified the facilitators and barriers to vaccine offer and uptake presented below, firstly from the perspective of GBMSM and secondly from the sexual health service provider perspective. These are categorised according to the constructs of the COM-B model of behaviour change (see Figure 1).

**Figure 1.**
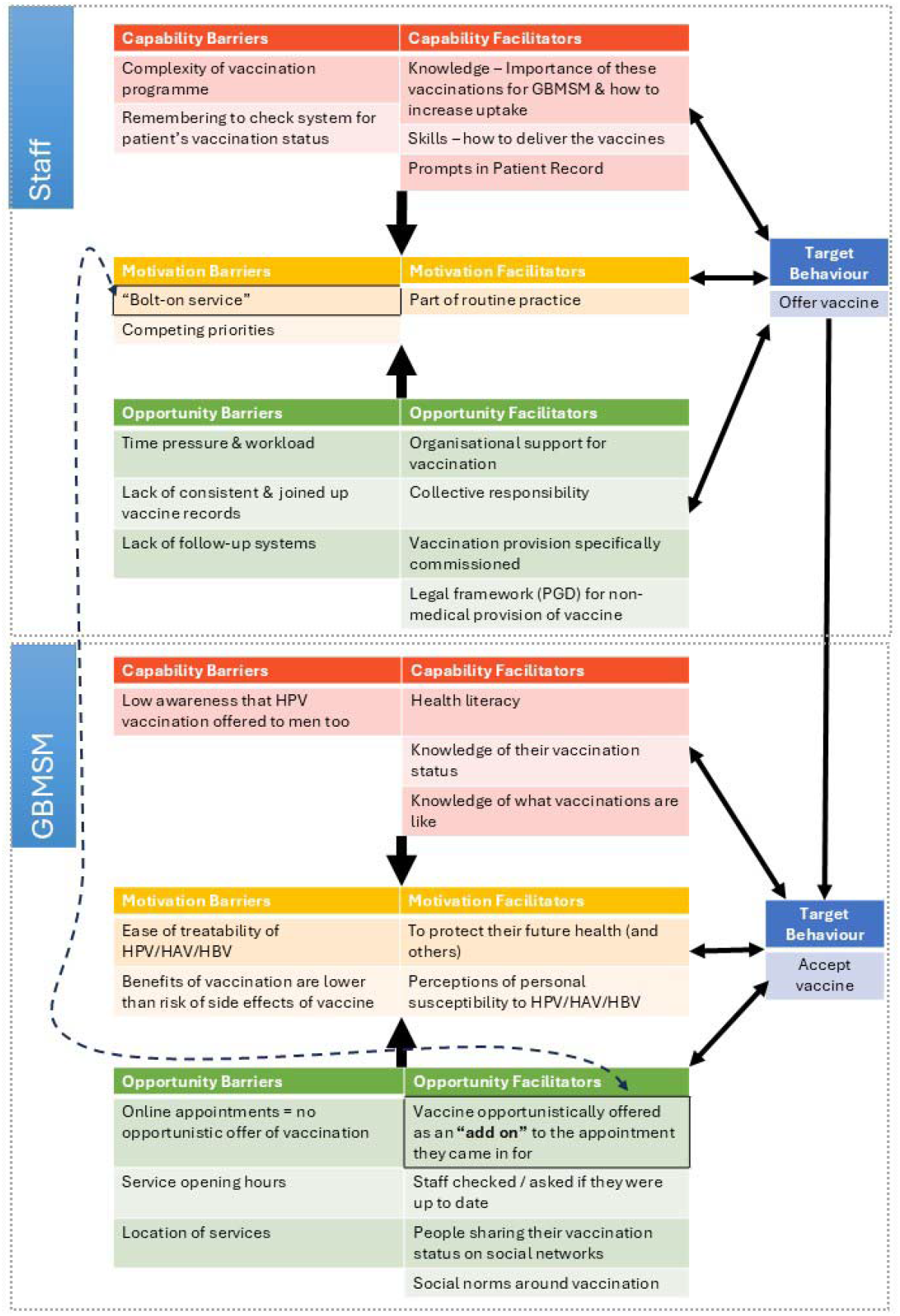
Diagram showing interacting COM-B components identified in the interview data.

### 3.1. Facilitators and barriers to vaccine uptake amongst GBMSM

#### 3.1.1. Capability

Most GBMSM participants who attended sexual health services described being proactive in maintaining their sexual health, which often supported uptake of opportunistic vaccine offers (See also 3.1.2. Opportunity). Although few said they had actively sought to be vaccinated, high-levels of health literacy and engagement with sexual health services meant that offers made during consultations were generally accepted:

> *I think it was offered to me […] as something that they offer free, not many side effects, like useful to have for my own protection. And I’m fine with vaccines, so I felt [it was] an easy yes, but I don’t think I’d seen anything about it before. It was just offered there and then* (GBMSM, South-West England, Interview 27)

Previous positive experiences with other vaccines and medications also reduced anxiety and facilitated uptake, with participants reporting any side effects as manageable and expected:

> *I wasn’t too worried really because I’ve experienced most usual side effects with medication and things like that anyway. So I know what it’s like. It is what it is* (GBMSM, North-West England, Interview 25)

In contrast, most of those with limited sexual health service engagement reported uncertainty, including a lack of knowledge about where vaccines could be accessed, and who was eligible to receive them:

> *I’ve never heard of the HPV vaccine, I haven’t heard of it before now [*…*] I don’t know if it’s being offered or if it’s being given anywhere* (GBMSM, South-West England, Interview 32)

Low vaccine awareness was often linked to assumptions that the HPV vaccination was intended exclusively for women or teenage girls:

> *The nurse was like, have you had the HPV vaccine and I was like, I thought that was totally for teenage girls, I didn’t know it was offered to anyone else* (GBMSM, North-West England, Interview 18)

#### 3.1.2. Opportunity

Engagement with online and offline GBMSM networks, including informal conversations with peers and the use of sexual networking apps where vaccination status was sometimes displayed, helped increase exposure to information and norms around vaccine availability:

> *I’d spoken to one of my friends, and I was like, ‘Oh, I’ve had my vaccinations and it opened up the broader conversation about sexual health, immunisations, and what people were doing* (GBMSM, North-West England, Interview 20)

Regular attendance at sexual health services for other needs, such as routine testing or PrEP appointments, further facilitated vaccination uptake. These consultations allowed staff to review vaccination status and either initiate or complete courses:

> *When I go in [every] three months for PrEP it’s like we just check all the vaccines are up to date and whether I need to have them again or when they need to be done* (GBMSM, North-West England, Interview 19)

For those attending sexual health services regularly, the convenience of receiving a vaccine during a routine appointment was more acceptable than having to seek it elsewhere. Preferences for combined injections (Twinrix) compared to separate HAV or HBV vaccines were also highlighted in one site:

> *There is a combined [vaccine] for A and B, but often we give Hep A, B, and HPV separately. Sometimes, they’re [patients] like, ‘Ooh, can I just have two today?’ or, ‘Can I just have one today?’* (Clinical Staff, South-West England, Interview 6)

Whilst connections to GBMSM networks and social media channels were found to increase exposure to vaccine-related information, those disengaged or marginalised from these networks reported having limited access to such information:

> *I’ve not gone to saunas, so I’d not experienced that […] So I’ve missed out on the catchment of people being there to provide me information* (GBMSM, North-West England, Interview 20)

Logistical issues - such as clinic opening hours and difficulties booking appointments - reduced access to in-person services. The increased use of online postal self-sampling kits for STI testing, introduced to mitigate these barriers, also reduced opportunities for in-person contact, thereby limiting chances for opportunistic vaccination:

> *It’s become really easy cos you can order the kit to your house, and you don’t have to interact with anybody anymore. So I think point of interaction is really reduced unless you have symptoms […] if it’s really difficult to get access to somebody to talk to they’re not going to do it* (GBMSM, South-West England, Interview 1)

#### 3.1.3. Motivation

Perceptions of personal susceptibility shaped how participants thought about vaccination. For some, engagement in sexual behaviours that were considered ‘high risk’ reinforced vaccination as an important form of protection:

> *For someone like me who is gay and I’m having sex sometimes – most of the time I’m bareback without condom, it’s important for this vaccination because we are very prone to STDs […] we are very exposed to hepatitis virus* (GBMSM, South-West England, Interview 26)

Vaccination was framed by some participants as a way to protect their overall health and maintain peace of mind or as part of a duty to safeguard partners and the wider LGBTQ+ community, given the disproportionate impact of STIs on GBMSM:

> *I feel a little bit like there’s a bit of a responsibility, as a gay man, to take seriously the fact that some of these viruses impact us disproportionately […] I wouldn’t want to put anybody at risk by virtue of the behaviours I choose to engage in* (GBMSM, North-West England, Interview 8)

Nevertheless, some participants who used PrEP reported that the protection afforded by medication against HIV led them to worry less about acquiring other infections such as HPV, which were perceived as less severe. In turn, this reduced the perceived importance of vaccination against them, as articulated by one participant:

> *HIV is just like the main STI and that that’s what we should focus on compared to other STIs […] that’s what I feel personally, and that’s why maybe I’m not really putting much effort to get about and getting a vaccine for HPV* (GBMSM, South-West England, Interview 32)

For some, low perceived severity of infections, combined with concerns about potential vaccination side effects, led them to decline vaccine offers in clinic. In these cases, the perceived benefits of vaccination did not always outweigh the perceived risks of side effects:

> *Before COVID I had every single vaccine that was going and I was annually vaccinated for flu and absolutely no problem at all. I had the two COVID vaccines back in 2021, but since the problems that have come out over vaccines since then I haven’t had a single vaccine […] HPV just doesn’t worry me at all [*…*] It’s at the back of my mind and I keep thinking ‘oh, I ought to have it’ but, as I said, every time they offer me the vaccine I’m worried about what the side effects might be and am I going to feel ill for a week after the vaccine or is it going to cause some kind of unexpected side effect like the COVID vaccine did on some people back in 2021* (GBMSM, North-West England, Interview 14)

### 3.2. Facilitators and barriers to vaccine programme delivery for service providers

#### 3.2.1. Capability

Sexual health service staff reported that they received training promoting the importance of vaccine delivery and how to encourage uptake. Staff felt knowledgeable enough to identify eligible patients, provide information and discuss the benefits and risks of vaccination with GBMSM. Prompts in electronic patient record proformas/templates also helped remind staff to offer patients vaccination (see also 3.2.2 Opportunity).

Nevertheless, some staff viewed the vaccination programme as complex and potentially confusing, given the varying number of doses, time points and schedule options (standard, rapid, and ultra-rapid) across vaccine types:

*If you’re started on one, do you carry on? What if you miss a dose? How then do you pick up on all the remaining doses?* (Clinical Staff, South-West England, Interview 12)

#### 3.2.2. Opportunity

Patient Group Directions (PGD) were in place to allow less senior staff to administer vaccines without needing sign off from a prescribing clinician. Flexibility in appointment length in sexual health services - facilitated by staff working from a shared patient list – also enabled vaccination and meeting other clinical needs:

> *[Appointments] they’ll always be much longer, but there is the expectation that some will be longer than others – and some will be quick and some will be not. We’ve now got [a] pooled central list that we all pick patients from – so you know that, if someone’s just stuck on someone, it doesn’t matter ‘cause someone else’ll pick up that patient*. (Clinical Staff, South-West England, Interview 6)

A sense of collective responsibility for offering, delivering, accepting and promoting vaccinations was also evident among commissioners and clinical teams (See also Motivation 3.2.3)

> *I think it [responsibility for GBMSM vaccination] sits with the system really. You could quite easily say ‘oh no, it sits with the clinicians; they are the ones who are seeing the individuals face to face’ but that’s not really true. We all have a part to play. Communications, marketing, health promotion, those are everyone’s responsibility* (Non-clinical Staff, North-West England, Interview 11)

To help reduce the risk of clinicians forgetting to offer vaccines, prompts in electronic patient record proformas/templates helped to support optimal delivery (See also Capability 3.2.1):

> *Our proformas are all set up so if you open a new patient pro forma it filters. If you enter in somebody’s GBMSM then it will automatically come up with questions asking you about their vaccination status. So there are prompts. I’m not saying that we always complete those but there are certainly prompts there to prompt us to do that* (Clinical Staff, North-West England, Interview 5)

Course completion was considered more challenging than delivering the first dose. This was partly attributed to capacity constraints and the need to remember to check vaccination records, as existing systems did not provide automatic reminders when subsequent vaccination was due and this information was not always easy to locate. Some staff described a ‘passive’ approach, whereby patients were expected to book their next dose themselves, although this relied on patients remembering, feeling confident to request vaccination or having another reason to re-attend. Staff recognised that this could lead to missed opportunities, as vaccination alone was not always perceived as a strong enough motivator for return visits:

> *Do we proactively book them an appointment to come back for their next vaccine? Or do we leave it up to them [*…*] But then that’s putting the emphasis and the onus on that individual, and whether or not they remember themselves, or feel comfortable asking, is of course, an entirely different matter* (Clinical Staff, South-West England, Interview 12)

The lack of open-access patient records to verify self-reported vaccination status across healthcare providers and settings (e.g. GP), and the lack of follow-up systems (e.g. automated reminders) to support course completion, also acted as barriers to vaccination. When vaccination records were incomplete or unavailable, staff used varied approaches in place of a consistent system, including estimating status based on age (e.g. comparing to introduction of the universal HPV vaccination programme), recall, previous attendance, or travel history (for HAV); asking patients to confirm details later; contacting other clinics; restarting vaccination courses; or continuing from the dose patients believed they had received. However, these processes were inconsistent within and across services:

> *We have no particular policy in place for chasing that information so that would be left with the clinician to make a decision. We could ask our health advisers to ring the other clinic and clarify what vaccinations they have had and that would be best practice but I’m not naive enough to believe that that happens every time* (Clinical Staff, North-West England, Interview 5)

Interviewees discussed potential ways to address these issues, including patient level vaccination records (e.g. online or physical records) and system wide changes supporting data sharing across services to enable clinicians to see what patients have received elsewhere.

Time and workload pressures - particularly in clinics with high patient volumes and complex patient needs - meant that data entry was often deprioritised in favour of direct clinical care. This had the potential to reduce the accuracy of surveillance data and compromised the usefulness of the data for supporting service improvement:

> *Practically speaking what that means is you’ve got a room full of people waiting and at the end of your half hour appointment you’ve then got to code before you see your next person, and the reality is if something is going to give it’s going to be the coding* (Clinical Staff, South-West England, Interview 7)

There was also a reflection that unless vaccination is part of local commissioning specification documents and costed into service models, organisational level support for providing vaccines may reduce:

> *In times of really tight budgets for commissioners, particularly local authorities, I think if you are not careful and you don’t put it in then you run the risk of being told by the provider ‘well you didn’t put that in the spec so we’re not doing it*’ (Non-clinical Staff, South-West England, Interview 21)

#### 3.2.3. Motivation

Staff felt confident to identify eligible patients and saw offering vaccines as a routine part of most consultations, following sexual history taking and addressing the reason for consulting.

In contrast to the suboptimal vaccination uptake estimated in England by national surveillance data, staff also described being generally diligent about completing patient records and surveillance systems with vaccine delivery data. Data entry was recognised as essential for monitoring vaccine uptake trends, identifying areas for service improvement, and securing funding. Examples were given of changes to services in response to data and regular reviews, training and individual and team feedback on data entry in one service:

> *Coding is how we get our funding, it’s how we monitor activity, it’s how we can pick up on trends and things like that* (Clinical Staff, South-West England, Interview 12)

Although staff were motivated to enter vaccination data, this was sometimes hindered by time and workload pressures caused by high patient volumes and complex patient needs (see also 3.2.2 Opportunity).

Provision of vaccines was not usually the primary focus of consultations and in one interview it was described as a *‘bolt-on’* activity (Staff interview 12), which may not be performed when clinical priorities or patient needs take precedence. Needing to prioritise more serious issues, managing complex consultations or avoiding overwhelming the patient meant vaccination was not always offered:

> *I mean vaccines usually comes last because you’re sometimes dealing with more immediate things. So there’s kind of a HIV risk now and we need to talk about PEP now, and that has to be done within 72 hours but before that we need a point of care test. So vaccines can be delayed a bit, you know, if it isn’t the right […] because what you don’t wanna do is scare them off* (Clinical staff, North-West, Interview 4)

## 4. Discussion

This study highlights the factors influencing the opportunistic offer and uptake of HAV, HBV and HPV vaccination among GBMSM within sexual health services. Consistent with earlier research demonstrating the acceptability of the HPV vaccine in this population (37, 38), our findings show that co-administered, opportunistic vaccines were generally acceptable to GBMSM, even among those with limited knowledge of HAV, HBV and HPV, or those unaware that vaccination is recommended. Vaccine delivery was also perceived as acceptable by staff working in clinical settings, although several system-level barriers limited optimal coverage. Additionally, the programme had limited reach beyond sexual health service settings, particularly among populations underserved by current opportunistic delivery models.

GBMSM appeared to have low knowledge of the three viruses, including symptom recognition, long-term health complications and the availability of vaccines through sexual health services. Previous research has documented that low knowledge and awareness of HPV, as well as vaccination as a method of prevention, is common among GBMSM (29, 37). Our findings extend this by highlighting similar gaps for HAV and HBV, including knowledge about vaccine entitlement and eligibility (psychological capability). Nevertheless, limited knowledge did not reduce willingness (reflective motivation) to receive these vaccinations, which was often supported by discussions with healthcare providers (facilitating psychological capability) and the convenience (physical opportunity) of receiving vaccines during routine sexual health visits. This aligns with previous work emphasizing the importance of healthcare provider recommendations and reminders (enhancing psychological capability), as attitudes (reflective motivation) toward vaccines are often influenced by the clinical environment and reassurance from trusted professionals (37, 38, 40). Hence, integrating vaccination within routine sexual health services should be viewed as an efficient and convenient (physical opportunity) strategy for GBMSM with regular engagement with sexual health services.

Such an approach, however, is only likely to be effective if vaccines are equitably accessible through these settings. Commonly, individuals received vaccines only after attending clinics for other healthcare appointments (e.g. PrEP), rather than actively seeking them (see also (37)). This was often due to limited awareness of vaccine eligibility (psychological capability barrier) and low perceived severity of the viruses (reflective motivation barrier). With increasing reliance on digital and remote service provision, opportunities for opportunistic vaccine delivery during in-person visits may decline (physical opportunity barrier), particularly for those individuals in this study with low severity perceptions of the viruses, potentially leading to suboptimal uptake (29). Additional efforts are therefore needed to increase knowledge of HPV, HAV and HBV, as well as awareness of vaccine eligibility. This includes addressing psychological barriers - such as beliefs that HPV is *“treatable”* or perceptions that vaccination is unnecessary - which reflect deficits in reflexive motivation and capability. Such efforts are particularly important among populations with lower levels of clinical engagement, including minoritized ethnic groups, young people and bisexual or straight-identifying MSM (10, 41).

Opportunities to obtain vaccine-related information varied depending on individuals’ engagement with both social and community networks and sexual health services. For many participants, exposure to information - including awareness that vaccines were available through sexual health services - occurred primarily through social connections to online and offline GBMSM networks (social opportunities facilitating increased psychological capability). These included informal conversations with peers, as well as sexual networking apps where vaccination status was sometimes displayed. Being socially connected within GBMSM communities has been associated with higher vaccination uptake in other contexts, such as during the 2022 mpox outbreak (42, 43), likely facilitated by the sharing of information about vaccination within social networks (39). In contrast, uptake of HPV, HBV and HCV vaccines appears to be lower among GBMSM with lower markers of social capital (10). Studies on HIV self-testing (44) and PrEP uptake (45) have similarly found that social and geographical marginalization from GBMSM networks can reduce individuals’ exposure to information about these interventions. Future vaccination campaigns should therefore target individuals marginalized from these networks to avoid reinforcing existing disparities in awareness, access and uptake. Here, targeted messaging emphasizing personal relevance and the benefits of vaccination should be used to improve awareness of personal risk and support informed decision-making (39), whilst community-led outreach and vaccination delivery through alternative routes - such as pharmacies or outreach vans - can help to reduce structural barriers to uptake (46–48).

A novel feature of this work was the inclusion of participants who had actively declined vaccination or had no intention of accepting it if offered. Some reported that their decisions were influenced by what they perceived to be side effects experienced following COVID-19 vaccination, leading them to avoid additional vaccines for viruses they perceived as ‘low risk’ (reflective motivation barriers). While this suggests a potential disconnect between perceived and actual risk among some GBMSM, it also reflects broader post-COVID skepticism toward vaccination, where concerns about side effects and exposure to multiple vaccines are shaping decision making around uptake (49). It should be noted that, in the broader GBMSM population, COVID-19 vaccination uptake has been reported as high (50), suggesting that these concerns may be concentrated in a small subgroup of the GBMSM population. Given the recent introduction of routine Mpox and gonorrhoea vaccination programmes for GBMSM in the UK, it is important to consider how additional vaccines may influence uptake of other vaccines targeting infections perceived as ‘low risk’ among GBMSM, including HAV, HBV and HPV (38), particularly if concerns about side effects or reactogenicity may affect willingness to complete vaccination schedules. Such insights can help guide the design and roll-out of future vaccination programmes by highlighting the factors that influence engagement and uptake within GBMSM communities.

Clinic staff were generally knowledgeable and confident in identifying eligible patients and promoting vaccines, supported by training and electronic record prompts. However, logistical and structural challenges - including complex vaccine schedules, inconsistent access to verified vaccination histories, high workload and reliance on patients to self-initiate return visits for follow-up doses - limited the consistency of vaccine delivery and course completion.

Given that clinician recommendations and clear guidance on follow-up doses are strong (motivational) facilitators of vaccine course completion (29, 38), additional measures are needed to optimize clinicians’ role in delivery and mitigate these system-level (physical opportunity) barriers. These include implementing more robust, joined up and consistent systems for patient vaccination status record access and patient follow-up, particularly when patients are unsure of their vaccination status or report receiving vaccines elsewhere at first offer. Systems to support course completion - such as contacting patients or using reminder mechanisms or prompts, ideally automated - would help ensure timely dosing. Introducing prompts for healthcare professionals (to address psychological capability barriers) could further support opportunistic vaccine completion during clinic visits for other reasons. Enhanced integration of vaccination within existing successful services could also be leveraged (supporting both psychological capability and physical opportunity): for example, aligning vaccine schedules with routine PrEP monitoring appointments may streamline delivery and reduce missed opportunities for follow-up doses, though differences in PrEP prescribing intervals, including 6 monthly reviews for some individuals, may limit this in practice. Finally, providing more feedback on service performance based on surveillance data reporting, (to support reflective motivation) and to allow services to check that what is reported is accurate (to facilitate psychological capability), could help optimise vaccination programme delivery.

The strengths of this research include exploring barriers and facilitators to all vaccines in the programme rather than focusing on individual vaccines in isolation, conducting interviews in two geographical areas with a diverse sample of GBMSM and staff holding a range of roles. Contextual differences at the clinic and system level between the two areas were identified which supported or restricted vaccination delivery. For instance, one area’s data system supported monitoring of vaccination through automated coding. Examining differences and similarities between GBMSM and staff perspectives illuminated areas for improvement with the programme. Without interviewing staff we would not have identified that course completion is a major issue or illuminated other clinic and system level issues. A limitation of the study was that despite employing a multi-pronged recruitment strategy including inside and outside clinics, our sample included only a small number of GBMSM with limited experience of sexual health services or those expressing vaccine hesitancy. However, we were able to indirectly capture these perspectives via staff accounts and GBMSM reflecting on their peers’ behaviour.

## 5. Conclusion

This study demonstrates that opportunistic delivery of HAV, HBV and HPV vaccines within sexual health services is mostly acceptable and feasible for GBMSM and for the staff responsible for administering vaccines. Despite low knowledge of these viruses and their associated risks, willingness to be vaccinated remained high, with healthcare provider recommendations and the convenience of vaccine delivery during routine clinic visits acting as important facilitators. However, the reach of opportunistic models was limited, particularly for individuals underserved by sexual health services or socially disengaged from GBMSM networks. System-level barriers such as complex vaccine schedules, inconsistent access to vaccination histories and reliance on patients to self-initiate follow-up doses were also found to contribute to sub-optimal vaccine coverage within services.

To improve equitable uptake, sexual health services should explore the feasibility of addressing both individual and structural barriers through additional strategies, including targeted and persuasive communication to increase knowledge, leveraging of regular contacts with patients and enhanced approaches to support vaccination completion (e.g. digital vaccine apps and prompts or reminders to support completion). Ensuring interventions are also attentive to the diverse needs, experiences and preferences of GBMSM - particularly those less connected to clinical or social networks - while promoting vaccination as a core component of sexual health care rather than an optional add-on, will also be key to enhancing acceptability, accessibility and equitable coverage.

As sexual health services now offer an increasing number of vaccines for GBMSM, the value and impact of these programmes will depend on achieving high and equitable coverage. The findings from this study, along with future work on the acceptability of new vaccination programmes, can help towards optimizing their delivery and coverage.

## Supplementary Materials

The interview topic guides can be downloaded at: https://www.mdpi.com/article/doi/s1

## Author Contributions

Conceptualization, CT, HF, JMK, TM, JH, LY, MC, SM, AW; methodology, CT, HF, JMK, TM, LY, JH; investigation, TM, JMK, formal analysis, TM, JMK, HEF; writing—original draft preparation, CT, TM, JK, HEF; writing—review and editing, TM, JMK, CT, HEF; HF, JH, AW, MC, HM, DL, LY, SM; supervision, CT, JH; project administration, CT, HF, TM, JMK; funding acquisition, CT, HF, JH. All authors have read and agreed to the published version of the manuscript.

## Funding

This study was funded by the NIHR Health Protection Research Unit in Behavioural Science and Evaluation at University of Bristol (NIHR200877), in partnership with UK Health Security Agency (UKHSA). The views expressed are those of the author and not necessarily those of the NIHR, the Department of Health and Social Care, or UKHSA

## Institutional Review Board Statement

The study was conducted in accordance with the Declaration of Helsinki, and approved by the UK National Health Service (NHS) Research Ethics Committee (REC reference: 23/YH/0159 dated: 14/9/2023) and the Health Research Authority (IRAS: 328525) prior to the study commencing.

## Informed Consent Statement

Informed consent was obtained from all subjects involved in the study.

## Data Availability Statement

Interview data will be available from the University of Bristol data repository, data.bris. Data access is restricted to bona fide researchers for ethically approved research and subject to approval by the University’s Data Access Committee.

## Acknowledgments

JMK, CT, HEF and JH are partly funded by NIHR Health Protection Research Unit in Evaluation and Behavioural Science at University of Bristol and NIHR Applied Research Collaboration West (NIHR ARC West). LY is an NIHR Senior Investigator and her research programme is partly supported by NIHR Applied Research Collaboration (ARC)-West and NIHR Health Protection Research Unit (HPRU) in Evaluation and Behavioural Science.

We are grateful for the contributions of GBMSM public contributors and study participants and participating sexual health service staff for their contribution to this study.

## Conflicts of Interest

The authors declare no conflicts of interest. The funders had no role in the design of the study; in the collection, analyses, or interpretation of data; in the writing of the manuscript; or in the decision to publish the results.

## Abbreviations

The following abbreviations are used in this manuscript:

GBMSM: Gay, Bisexual or other Men who have Sex with Men
GP: General Practitioner
HAV: Hepatitis A Virus
HBV: Hepatitis B Virus
HIV: Human Immunodeficiency Virus
HPV: Human Papillomavirus
LGBTQ+: Lesbian, Gay, Bisexual, Transgender, Queer, plus
NHS: National Health Service
NIHR: National Institute of Health and Care Research
PBA: Person Based Approach
PrEP: Pre-exposure Prophylaxis
STI: Sexually Transmitted Infection
UK: United Kingdom
UKHSA: UK Health Security Agency

## Disclaimer/Publisher’s Note

The statements, opinions and data contained in all publications are solely those of the individual author(s) and contributor(s) and not of MDPI and/or the editor(s). MDPI and/or the editor(s) disclaim responsibility for any injury to people or property resulting from any ideas, methods, instructions or products referred to in the content.

## Footnotes

1 Although universal vaccination programmes have been introduced for HPV (initially for girls in 2008 and extended to boys in 2019) and HBV (from 2017), many adults currently attending sexual health services - particularly GBMSM - will not have been fully covered by these programmes, and there is no universal vaccination programme for HAV.

2 An Intervention Planning Table is a tool commonly used in the early stages of intervention development, whereby PPIE discussions, relevant literature and qualitative data are collated to identify context-specific behavioural issues around the target behaviour(s) and potential intervention elements.

## Notes

### Competing Interest Statement

The authors have declared no competing interest.

