## Supplementary material for "Understanding Vaccination Uptake amongst Gay, Bisexual and other Men who have Sex with Men in UK Sexual Health Services: A Qualitative Interview Study": Supplementary Materials_Participant Topic Guide Interview.docx

**Explain purpose of project and session**

NHS guidelines recommend that HPV vaccination be offered to GBMSM up to and including age 45 years when they attend sexual health services (SHS) and HIV clinics. Guidance also promotes the offer of Hep A (HAV) and Hep B (HBV) vaccination to GBMSM of any age in the same settings.

We are trying to find out from Gay, Bisexual or other Men who have sex with men what concerns they may have about vaccines offered in sexual health clinics. We would also like to design new communication materials which can be used to help individuals make informed decisions about the recommended vaccines. We anticipate that the interview will last up to one hour.

We are also talking to professionals involved in delivery of the vaccination programmes in sexual health clinics.

[Note: *Ensure that the participant has read the information sheet and completed the the consent form electronically or provided verbal consent before the interview starts*.]

### (1) HPV/HAV/HBV awareness and perceptions

Can you tell me what you know about:

- Human Papillomavirus (also called HPV)
- Hepatitis A
- Hepatitis B

Prompts: Symptoms? Routes of transmission? Diseases they can cause? Prevention

What are the risks of contracting HPV/HAV/HBV?

Provide brief information to participant if required: Persistent infection with HPV can lead to the development of anogenital warts and oropharyngeal, genital, and anal cancers. Infections with HAV and HBV can lead to liver damage, cancer, or failure.

How at risk do you think you are from acquiring:

- HPV
- Hepatitis A
- Hepatitis B

Do these risk perceptions differ by virus? Why is that?

How serious do you think it would be if you had an infection with any of these?

What risk to others do you think you might pose if you had any of these infections?

Do you think any of these viruses are more severe than the others?

- If so, why?

### (2) Vaccine decision-making

Have you received vaccination for HPV/HAV/HBV? (or currently receiving a vaccination course?)

**IF YES**

- What have you been vaccinated for?
- Have you completed the full course?
- What was the most important reason for being vaccinated?
  - Protect others including family and friends, the community, oneself?
  - Influence of outbreaks on attitudes towards vaccination among clinicians and patients (e.g. COVID, mpox)
- Did you have any worries about receiving the vaccine?
- Where did you receive the vaccine(s)? Was it accessible in terms of location/time/opening hours?
- Can you tell me a little bit about the process?
  - Ask to explain procedures – were they invited to come and have the vaccine, was it offered immediately, do they have to go back for further doses? Are there any difficulties and challenges in GBMSM completing the full course of each vaccination at different timepoints?

*Vaccine information*

How did you find out about the vaccine?

- Word of mouth, email or text message from sexual health clinic, recommendation from professional during sexual health appointment, other, posters / social media campaigns?

What information were you given and by whom? What did you think about this information (trust, answer their questions, easy to understand)?

Who did you talk to about the vaccine?

What other information did you look for?

**IF NO OR NOT RECEIVED ALL VACCINES/DOSES**

- Can you tell me the most important reason why you have not had the vaccine or completed the schedule?
- How would you feel about being vaccinated against HPV/HAV/HBV? Do you think it is important?
- How willing are you to receive any of these vaccines?
- Do you think the vaccine(s) is/are safe and effective?

Do you know where you could receive a vaccine? Would you know how to book/ask about a vaccine(s)?

- How do you feel about being vaccinated in a sexual health clinic? Do you think this is the best place for GBMSM to receive vaccinations?
  - Prompts: Is it accessible (location and operating hours?), do you have time to go and get vaccinated?

**FOR ALL**

Do you think most within the GBMSM community have had or will plan to have the vaccine(s)? Is this seen as being the ‘norm’ or ‘the right thing to do’?

Do you know any others, including influential people from the GBMSM community or friends/peers, who have received the vaccine(s)?

For the GBMSM community, what do you think might be the main difficulties and challenges in getting these vaccines?

- Knowledge and awareness of availability of vaccine
- Perceptions of need
- Perceptions of severity
- Perceptions of susceptibility
- Perceptions of benefits
- Stigma of being vaccinated
- Logistics/acceptability of SHS as settings
- Trust in public health programmes
- Government intrusion in private lives
- Misinformation or information known to be fake or not trusted
- Perceived vaccine efficacy

**Comparison to national data**

*Explain to the patient that we have been provided with national level summary statistics of HPV, Hep A and Hep B vaccination uptake among MSM who attend sexual health clinics for an appointment.*

*[N.B. Further data available in summary document within data* *folder**]*

*Ask the patient to comment on whether the following data aligns with their perception of who might be more likely to take up vaccination among MSM community..*

- Post-pandemic, online consultations have continued to rise. Do you know if your local sexual health clinic offers online or telephone consultations?
- 21% uptake of first dose for HPV, reducing to 8% and 7% for subsequent doses.
- 10% uptake of first dose for Hep B, reducing to 8% and 5%
- 6% uptake of first dose for Hep A, reducing to 3% for second dose
- Younger cohorts more likely to be vaccinated than older cohorts
- Known HIV positive status less likely to be vaccinated – ask about clinicians working across Brecon Unit and Unity.
- No differences apparent between urban and rural settings
- We cannot be confident in the data on ethnicity and socio-economic status because the levels of missing data mean they may not represent an accurate picture of any differences in uptake associated with these factors.

3) **Developing communication materials**

*As part of this study, we are hoping to design new communication materials to complement the existing leaflet produced for vaccination programmes delivered in sexual health clinics.*

What format do you think the new materials should be?

- Videos
- Leaflets
- Web pages
- What information do you think are the most important things to include?
- Free, confidential, quick etc
- How HPV/Hepatitis A/Hepatitis B are spread
- Diseases that HPV/Hepatitis A/Hepatitis B can cause
- Need for vaccination (e.g. not sexually active, cultural and religious beliefs)
- Target age when vaccination offered
- Safety
- Side-effects
- Research into vaccine
- Anything else?

Where should information come from?

- Media campaigns
- Health fairs / events
- Community advocates / vaccine champions
- General practices
- Pharmacists
- Dental settings
- One-to-one with healthcare professionals
- Web-based forums
- Interactive information sessions
- Trusted GBMSM sources/messengers – please provide examples

Whose responsibility do you think it is to make sure GBMSM are vaccinated?

- Sexual health service providers
- GP
- GBMSM community responsibility
- Other healthcare professionals

What could we do to make it easier for GBMSM to get the vaccines?

- Incentives
- Pharmacy led clinics
- Social media campaigns
- Community out reach
- Reminders for vaccination and follow-up doses
- Vaccination clinics
- Creation of a GBMSM schedule similar to childhood vaccination (including mpox, menB as well as HPV, HBV, HAV)
- Help navigating multiple doses of vaccines

**Finally, is there anything else you would like to tell me or ask me about?**

**Many thanks for taking part in this interview.**

[NOTE: *ensure that the participant is given the £30 gift voucher and reimburse travel expenses*]
