## Supplementary material for "Understanding Vaccination Uptake amongst Gay, Bisexual and other Men who have Sex with Men in UK Sexual Health Services: A Qualitative Interview Study": Supplementary Materials_Professional Interview Topic Guide.docx

**Professional Topic Guide**

**Explain purpose of project and interview**

[Note: *Ensure that the participant has read the information sheet and completed the consent form electronically or provided verbal consent before the interview starts*.]

(1) **Role**

Please could you describe your role?

### (2) Delivery of vaccines

Can you tell me what you know about HPV/HAV/HBV vaccination for GBMSM?

Are these vaccinations available to GBMSM in your service or elsewhere? Since when? If not, why not?

How successful do you think the vaccination programmes are in your setting?

Whose responsibility do you think it is to make sure GBMSM are vaccinated?

- Sexual health service providers
- GP
- GBMSM community responsibility
- Other healthcare professionals

How is information about these vaccinations communicated to GBMSM?

- Information campaigns, posters in waiting rooms etc
- Letter of invitation
- Verbally by clinic staff, nurses, doctors etc
- Other community partners or services
- Email, text messages – move toward paperless approach
- Social media campaigns
- Posters

Do any communications/materials/protocols to promote vaccination in GBMSM exist within your service? Would you be willing to share these?

Can you tell me how vaccines are offered to GBMSM who attend SHS?

- Who is responsible for overseeing delivery of the vaccination programmes?
- Who is responsible for providing the vaccinations?
- Do you have vaccinations in stock on site?
- How do you identify who is eligible for vaccination? (clinical record search, questionnaire during appointment, pop-ups or flags in the record system)

What is the procedure for offering these vaccines to GBMSM?

- As part of routine appointments?
- For people who ask or self-identify as at risk?

Is there a system which is used to record or identify vaccinations status? What is it called?

- How is coding for vaccination completed on patient records and GUMCAD?
- What do people code for in GUMCAD?
- How do you code for a patient who has been vaccinated previously? For example, Hep A could be received as part of travel vaccinations
- What are the reasons for not completing coding? (issues with software, codes not available, time)
- What could improve coding? (software changes to prevent closing window until coding has been completed, incentivisation)
- How is vaccination course completion monitored (e.g. avoiding multiple first doses)?

### (3) Barriers and facilitators to uptake

For providers who deliver the vaccine, are there any difficulties and challenges in offering the vaccine to GBMSM?

- Lack of provider training and skills
- Lack of provider knowledge and awareness e.g. around eligibility criteria
- Forgetfulness
- Difficult to discuss
- Time constraints
- Perceptions of patients (low risk or low priority awareness of risk factors, perceived susceptibility of different viruses, perceived severity, perceived benefits, perceived barriers among different populations/demographic characteristics)
- Perception of clinicians (awareness of risk factors, perceived susceptibility to virus, perceived severity of virus, perceived benefits of offering vaccination)
- Differences in attitudes towards vaccination across patient demographic characteristics (such as ethnicity and age)
- Beliefs about offering (could offend patients by assuming sexual activity/promiscuity)
- Not the norm within consultations
- Provider-patient relationship (new patient, concern about privacy and confidentiality if patient is known to provider or family member present)
- Influence of outbreaks on attitudes towards vaccination among clinicians and patients (COVID, mpox)

For providers who deliver the vaccine, are there any difficulties and challenges in GBMSM completing the full course of each vaccination? (different timings of each vaccination)

- How do patients and clinicians navigate multiple doses of vaccines at different timepoints for each vaccine? (reminder systems)

What things help to motivate offering vaccinations?

What things help to motivate patients accepting vaccinations?

Can you suggest any changes within the clinic that might help overcome the barriers identified above and improve uptake?

- Use of pop-ups or reminders in the electronic record
- Availability of information resources to give to eligible patients
- Training / educational resources/awareness raising amongst staff
- Clearer guidelines or clinic protocols/procedures
- Online information / information outside the clinic motivating vaccination
- Reminders for vaccination and follow-up doses
- Vaccination clinics
- Creation of a GBMSM schedule similar to childhood vaccination (including mpox, MenB as well as HPV, HBV, HAV)
- What would help patients and staff navigate multiple doses of vaccines?

(4) **Comparison to national data**

*Explain to the provider that UKHSA have provided national level summary statistics of HPV, Hep A and Hep B vaccination uptake among MSM who attend sexual health clinics for an appointment. Clarify that this may not align with the figures for their own organisation.*

*[N.B. this section was not used with all participants]*

*Ask the provider to comment on whether the following data aligns with their perception of uptake within clinic.*

- Post-pandemic, online consultations have continued to rise. Does your clinic conduct online or telephone consultations or could GUMCAD data reflect online testing data?
- Do certain populations favour certain appointment types?
- Do you see some populations in person more (e.g. PrEP users) which could explain changes/differences in appointment types?
- 21% uptake of first dose for HPV, reducing to 8% and 7% for subsequent doses.
- 10% uptake of first dose for Hep B, reducing to 8% and 5%
- 6% uptake of first dose for Hep A, reducing to 3% for second dose
- Younger cohorts more likely to be vaccinated than older cohorts
- Patients with known HIV positive status less likely to be vaccinated
- No differences apparent between urban and rural settings
- We cannot be confident in the data on ethnicity and socio-economic status because the levels of missing data mean they may not represent an accurate picture of any differences in uptake associated with these factors.

**Finally, is there anything else you would like to tell me or ask me about?**

**Many thanks for taking part in this interview.**

[NOTE: *ensure that the participant is given the £30 gift voucher and reimburse travel expenses*]
